# From Paper Letters to an Integrated Digital Workflow: Improving Efficiency, Reliability, and Engagement in Health Guidance

**DOI:** 10.64898/2026.06.10.26355234

**Authors:** Ikumi Kakizaki, Emi Hirafuji, Midori Araba, Ryoko Yoshida, Yoichiro Aoki

**Author notes:** Corresponding author: Yoichiro Aoki.

## Abstract

**Background:** Post-checkup health guidance in Japan has traditionally relied on paper-based communication and manual administrative processes. These workflows are time-consuming, prone to transcription errors, and can delay timely engagement with health guidance recipients.

**Objective:** To assess whether replacing a paper-based workflow with an integrated digital system using Microsoft Access, robotic process automation (RPA), and web-based responses could improve administrative efficiency, operational reliability, and engagement among health guidance recipients.

**Methods:** This single-site quality improvement initiative redesigned the existing letter-based workflow. Access served as a central interface for managing recipients and generating guidance letters. RPA (EzRobot) automated repetitive clerical and billing-related tasks. A web form accessed via a QR code enabled recipients to respond digitally. Outcomes included manual administrative handling time per case, occurrence of transcription-related errors, health guidance completion rate, and guidance duration distribution.

**Results:** Following implementation, staff active handling time per case decreased from approximately 10 minutes to less than 1 minute (approximately 30 seconds), while automated RPA execution typically required about 4–5 minutes per case without staff input. No transcription-related errors were detected during the post-implementation observation period. Health guidance completion rates improved from 28.3% to 39.2% (χ^2^ test, P<0.01; R4 (FY2022) n=184, R5 (FY2023) n=536). Guidance duration distributions, calculated using the corrected method, shifted towards shorter durations: cases with ≥200 days decreased from 30.5% to 20.9% and cases with ≥240 days decreased from 13.6% to 8.9% (R4 n=59, R5 n=158).

**Conclusion:** An integrated Access–RPA–Web workflow was associated with improvements in administrative efficiency and operational reliability in post-checkup health guidance while retaining human verification and exception handling. This pragmatic, non-AI-dependent approach may offer a useful model for process-level improvement in preventive care settings.

**Summary Box (Key Messages):** *What is already known on this topic:* Paper-based health guidance remains widely used in preventive care, but it involves multiple manual steps, increasing administrative burden, delays in follow-up, and transcription risks. Robotic process automation (RPA) has been applied to healthcare administration, particularly billing and data entry, yet integrated workflows linking health guidance delivery, recipient responses, and billing transfer have rarely been described.

*What this study adds:* This study describes a non-AI-dependent digital workflow using Microsoft Access, rule-based RPA, and an optional QR-linked web form to modernise letter-based health guidance while retaining human verification and exception handling. The approach reduced administrative handling, automated routine transcription steps in billing transfer, and supported more efficient workflow management in routine practice.

*How this study might affect research, practice or policy:* Pragmatic digital integration may support sustainable quality improvement in preventive health services without major infrastructure investment. These principles may be transferable to other settings where paper-based communication remains necessary.

## Introduction

### Problem description

Routine health checkups are a cornerstone of preventive healthcare in Japan. Individuals identified as requiring follow-up guidance may be contacted by telephone or email, and paper letters are also used for ongoing support, followed by manual documentation and billing processes. Despite their widespread use, these paper-based workflows introduce several challenges: delayed initiation of guidance, high clerical burden for staff, variability in operational quality, and increased risk of human error during data transcription.^1,2^

### Available knowledge

Previous quality improvement and health informatics studies have demonstrated the benefits of RPA in healthcare administration, particularly in billing and registration processes. However, these studies largely focus on isolated administrative tasks rather than end-to-end guidance workflows. To date, there is limited evidence on integrated systems that modernise letter-based health guidance by combining database-driven management, administrative automation, and digital recipient responses within a single operational framework.^3,4^

### Rationale

Standardising the letter-based workflow through a unified database interface, automating deterministic clerical steps with rule-based RPA, and enabling recipients to respond via web forms address each of the identified failure points sequentially. This approach avoids the complexity and maintenance burden of artificial intelligence whilst offering meaningful improvements in safety and efficiency.

### Specific aims

This study aimed to evaluate whether transitioning from a conventional paper-based letter system to an integrated digital workflow using Microsoft Access, RPA, and web forms improves administrative efficiency, operational reliability, and engagement among health guidance recipients at a single preventive medicine centre in Japan.

## Methods

### Context

This quality improvement initiative was conducted at the Health Support Office, Preventive Medicine Centre, Yoshida Hospital, Keiyukai Medical Corporation, Japan. The centre provides routine health check-up services and associated health guidance to adults identified as requiring lifestyle modification support. Letter-based guidance is the primary method of ongoing contact with recipients. The Access database was limited to individuals eligible for health guidance; it did not include patients receiving ongoing outpatient clinical treatment at the hospital.

### Implementation (Workflow redesign)

The implementation replaced the existing paper-based workflow with an integrated digital process comprising three interdependent components (Figure 1).

**Figure 1.**
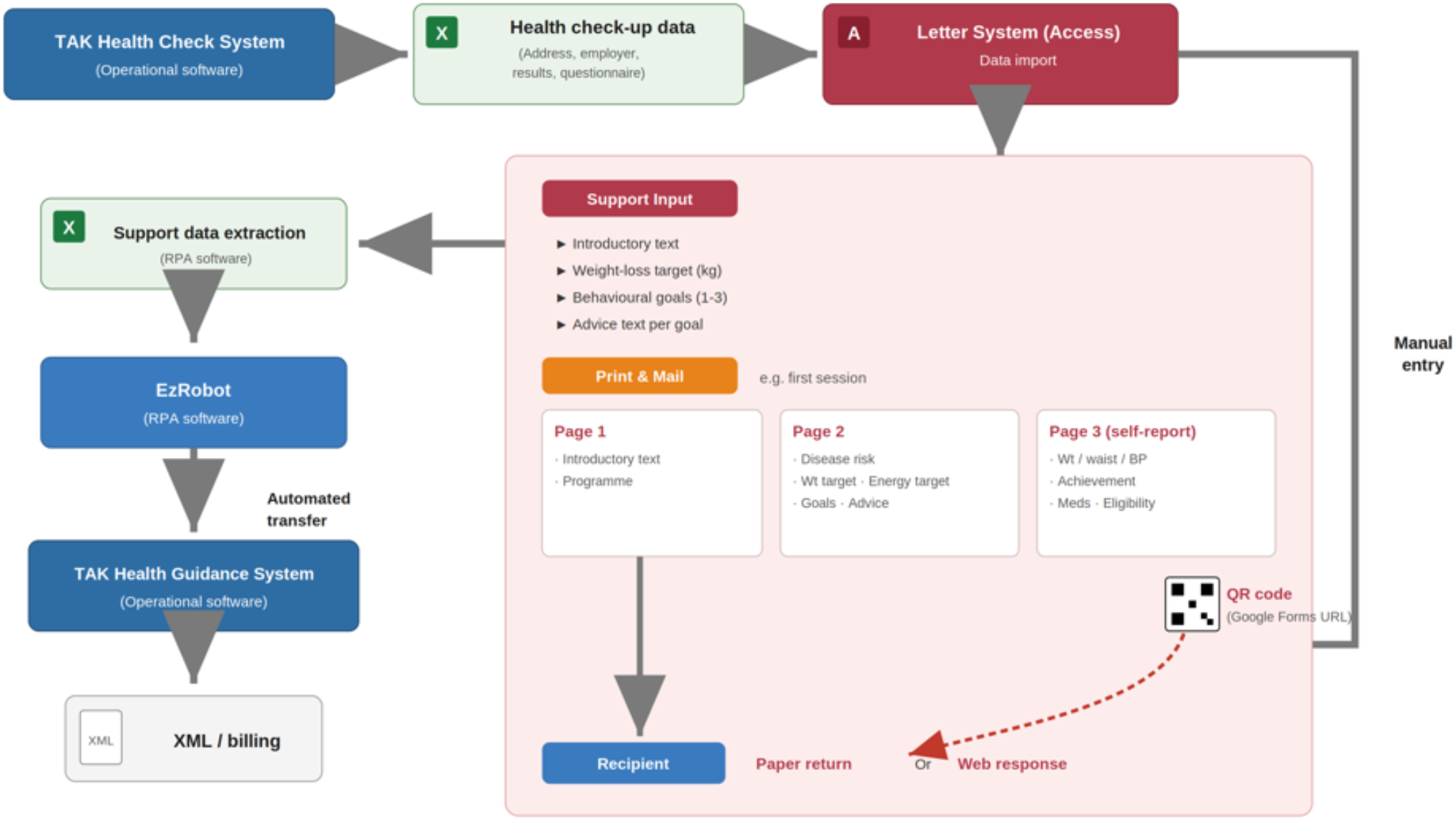
Integrated workflow for letter-based health guidance (Access database, robotic process automation (RPA), and optional web-based response form). Health check-up results, guidance content, and questionnaire data are centrally managed using a Microsoft Access–based system. Health professionals manually enter personalised guidance goals and messages based on clinical judgement. Rule-based RPA (EzRobot) is then used to automate predefined administrative tasks, including data extraction and transfer of required billing information into the health guidance management system. RPA is limited to deterministic, non-AI operations and does not generate or modify clinical content. Human verification and exception handling are retained throughout. To improve timeliness and engagement, recipients can respond either by returning paper questionnaires or by accessing a web-based response form via a QR code included in the guidance letter. Abbreviations: BP, blood pressure; Meds, medications; RPA, robotic process automation; Wt, weight; XML, Extensible Markup Language.

#### 1. Microsoft Access–based management interface

Microsoft Access functioned as a unified interface enabling staff to manage recipient lists, review health check-up results and questionnaire data, generate standardised guidance letters, track guidance status and timelines, and import digital responses from web forms. This consolidation replaced multiple spreadsheets and manual tracking methods, reducing variability in how staff managed cases and providing a single point of reference for all guidance activities.

#### 2. Rule-based robotic process automation

RPA software (EzRobot) was configured to automate repetitive clerical steps including Excel file processing, preparation of mailing and tracking lists, and the transfer of required billing data into the health guidance billing system. RPA was restricted to deterministic, non-AI operations and did not generate or modify clinical content. Staff retained full responsibility for clinical judgement and guidance decisions; only rule-defined clerical steps were automated. Human verification and exception handling were retained throughout the process.

#### 3. Web-based response system

Each guidance letter included a QR code linking to a designated web-based response form. Recipients could submit required questionnaire responses digitally, without returning paper documents. Submitted responses were automatically imported into the Access database, enabling rapid follow-up. The conventional paper return route was retained as a parallel option to preserve accessibility for all recipient groups.

### Study of the implementation

This was a before-and-after quality improvement evaluation at a single site. The conventional pre-implementation workflow was compared with the integrated digital workflow after phased implementation. Outcomes were compared between two consecutive Japanese fiscal years: Reiwa 4 (R4; fiscal year 2022, April 2022 to March 2023) and Reiwa 5 (R5; fiscal year 2023, April 2023 to March 2024). Implementation proceeded in three stages: first, workflow documentation and standardisation; second, Access database development and deployment; and third, RPA configuration and QR code integration.

### Measures

**Efficiency:** administrative processing time per case (minutes), defined as staff manual handling time required to complete clerical and billing-transfer tasks for one case.

**Reliability:** occurrence of transcription-related errors in billing transfer.

**Timeliness:** guidance duration distribution (≤160, 160–200, 200–240, ≥240 days) based on the corrected calculation method.

**Engagement:** health guidance completion rate (%).

### Analysis

Descriptive before-and-after comparison was used. Process measures were derived from operational records. Completion rates and the proportion of prolonged cases were calculated from guidance tracking data. For proportion outcomes, including completion rate and guidance duration categories, differences between periods were assessed using chi-square tests, and P values are reported where applicable. Other measures are presented descriptively in keeping with the pragmatic quality improvement design.

### Ethical considerations

This work was conducted as a quality improvement initiative using routine operational records. It did not involve experimental procedures, recruitment of participants, or automated clinical decision-making. Ethics approval was obtained from the Yoshida Hospital Ethics Committee, Keiyukai Medical Corporation, on 21 April 2026 (approval number: 20260421001). Individual informed consent was waived by the ethics committee because this quality improvement study used aggregated, anonymised operational data and posed minimal risk to participants. This report presents only aggregated results without identifiable personal information. Consent for publication: not applicable.

### Patient and public involvement

Patients and the public were not involved in the design, conduct, reporting, or dissemination plans of this quality improvement study. The study used aggregated, anonymised operational data and did not involve recruitment of participants.

## Results

### Implementation (Workflow redesign)

The integrated workflow was implemented in phases over the development period. Workflow standardisation and Access database deployment preceded RPA configuration, ensuring that process variability was reduced before automation was introduced. The QR code–linked web form was integrated in the final phase. Frontline staff were involved throughout, and the system was adapted iteratively based on operational feedback.

### Process measures and outcomes

Table 1 presents a comparison of key process measures before and after implementation. The detailed pre- and post-implementation operational indicators, denominators, and calculation methods are shown in Supplementary Table S1.

**Table 1.**
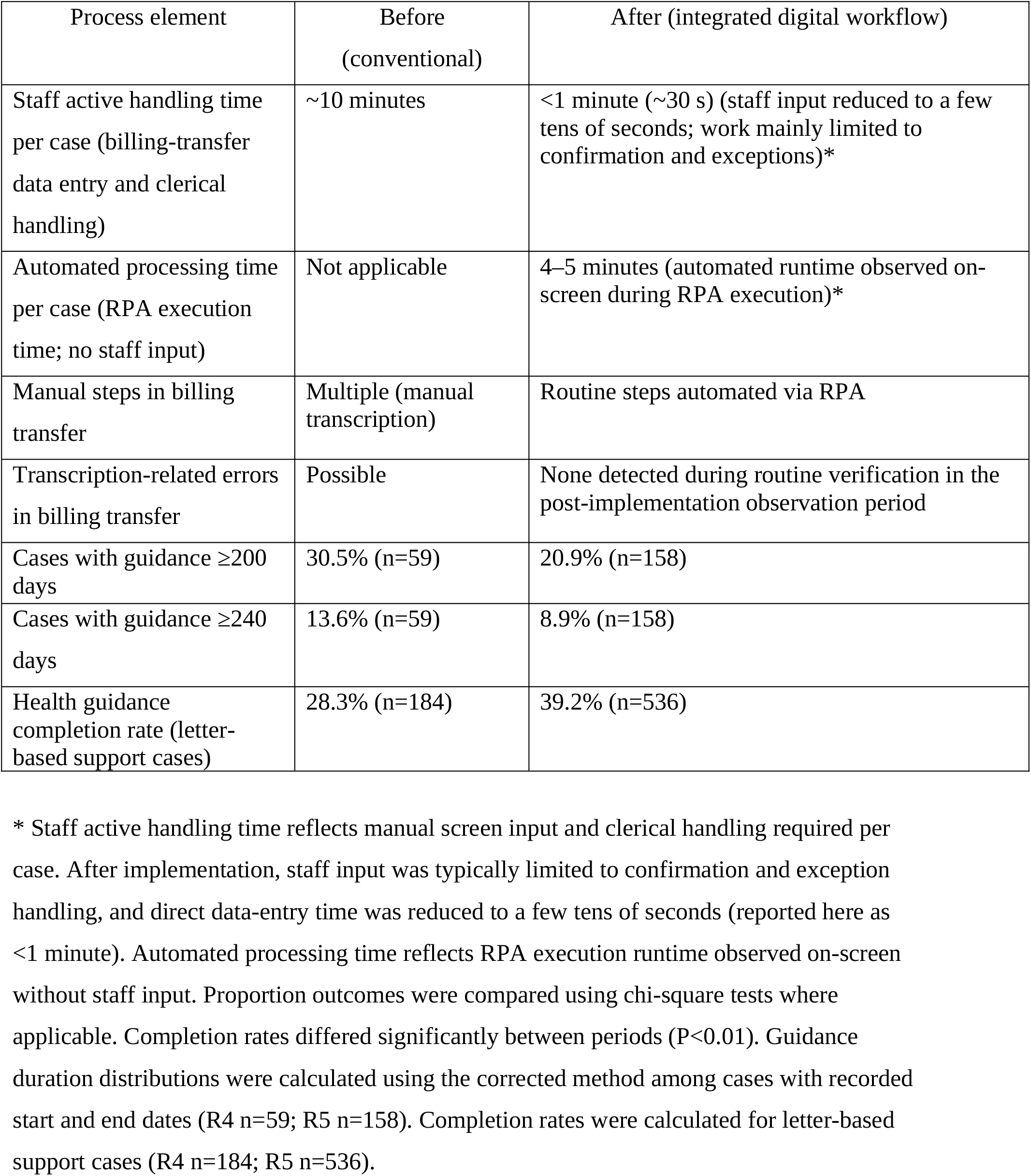
Before and after comparison of the health guidance workflow.

| Process element | Before<br>(conventional) | After (integrated digital workflow) |
| --- | --- | --- |
| Staff active handling time per case (billing-transfer data entry and clerical handling) | ~10 minutes | <1 minute (~30 s) (staff input reduced to a few tens of seconds; work mainly limited to confirmation and exceptions)* |
| Automated processing time per case (RPA execution time; no staff input) | Not applicable | 4–5 minutes (automated runtime observed on-screen during RPA execution)* |
| Manual steps in billing transfer | Multiple (manual transcription) | Routine steps automated via RPA |
| Transcription-related errors in billing transfer | Possible | None detected during routine verification in the post-implementation observation period |
| Cases with guidance $\geq 200$ days | 30.5% (n=59) | 20.9% (n=158) |
| Cases with guidance $\geq 240$ days | 13.6% (n=59) | 8.9% (n=158) |
| Health guidance completion rate (letter-based support cases) | 28.3% (n=184) | 39.2% (n=536) |

\* Staff active handling time reflects manual screen input and clerical handling required per case. After implementation, staff input was typically limited to confirmation and exception handling, and direct data-entry time was reduced to a few tens of seconds (reported here as <1 minute). Automated processing time reflects RPA execution runtime observed on-screen without staff input. Proportion outcomes were compared using chi-square tests where applicable. Completion rates differed significantly between periods ( $P < 0.01$ ). Guidance duration distributions were calculated using the corrected method among cases with recorded start and end dates (R4 n=59; R5 n=158). Completion rates were calculated for letter-based support cases (R4 n=184; R5 n=536).

To avoid implying a single nested cohort, the denominator for each outcome is reported separately, because the two outcomes were measured on differently defined operational subgroups. The engagement outcome (health-guidance completion rate) was calculated for all letter-based support cases, defined as recipients of mobile (roving) health guidance who were classified as requiring active support and who selected letter-based support at the second initial interview (R4 n=184; R5 n=536). The guidance-duration distribution was calculated for a separate subgroup, namely cases whose final evaluation was conducted by letter (R4 n=59; R5 n=158), drawn from the intensive-support pathway (eligible for intensive support: R4 n=756, R5 n=1,305; programme completed: R4 n=396, R5 n=674; Supplementary Table S1). Because these subgroups were defined on different operational bases, the completion-rate and guidance-duration denominators are not directly comparable and are presented as separate process measures rather than as a single funnel.

### Efficiency

Staff active handling time per case decreased from approximately 10 minutes to <1 minute (∼30 s) following automation of clerical and billing-transfer steps. Before implementation, staff typically spent about 10 minutes per case on manual screen input and administrative processing. After implementation, staff input was usually limited to confirmation and exception handling, and direct screen input was reduced to a few tens of seconds (reported here as <1 minute). The automated RPA execution itself typically required about 4–5 minutes per case on-screen, but this runtime did not require staff input.

### Reliability and timeliness

Automation removed manual transcription from the routine billing-transfer process. No transcription-related errors were detected during routine verification in the post-implementation observation period. Because the pre-implementation transcription-error rate had not been systematically recorded, this comparison is descriptive rather than quantitative. Using the corrected calculation method, longer-duration cases decreased (≥200 days: 30.5% to 20.9%; ≥240 days: 13.6% to 8.9%), consistent with improved timeliness of follow-up following implementation of the integrated workflow.

### Engagement

The health guidance completion rate increased from 28.3% to 39.2% (χ^2^ test, P<0.01) after implementation. The QR code–linked web form provided an additional response pathway for recipients and may have contributed to earlier engagement and more timely progression of cases.

### Contextual factors

Several contextual factors supported implementation. A designated staff member with both operational knowledge and practical information technology skills served as a bridge between workflow requirements and system design, enabling iterative refinement based on frontline experience. Organisational support for the initiative, including temporary redistribution of workload during implementation, and a receptive institutional culture towards healthcare digitalisation, facilitated adoption and sustained operation.

## Discussion

### Summary

This quality improvement report describes the design, implementation, and evaluation of an integrated digital workflow for letter-based health guidance at a single preventive medicine centre in Japan. The combination of Microsoft Access–based data management, rule-based RPA, and a QR code–linked web form was associated with greater administrative efficiency, improved timeliness of follow-up, and improved engagement as reflected by higher completion rates, while retaining human verification and exception handling throughout.

### Interpretation

The observed improvements were not merely a result of introducing new tools; several enabling factors also appeared to be important. First, a clear and persistent problem awareness existed within the frontline team. The existing workflow required substantial time for drafting, printing, envelope preparation, mailing, and handling responses, which contributed to delayed delivery and increased administrative burden. This shared recognition provided sustained motivation for improvement.

Second, the phased implementation approach was pivotal. The team did not begin with automation as the primary objective; instead, the letter-support workflow was first organised and standardised through the Access interface, and only then were routine tasks automated using RPA. This sequencing reduced process variation before automation was applied, making subsequent automation more reliable and easier to sustain.

Third, by restricting RPA to administrative and billing-related processes and retaining human confirmation and exception handling, the intervention improved operational efficiency without shifting clinical decision-making to automation. The non-AI, rule-based approach reduced maintenance burden and avoided concerns about automated clinical content generation.

### Limitations

This report has several limitations. First, it reflects a single-site implementation within a specific operational and policy context in Japan; generalisability to other settings requires consideration of local workflow, system infrastructure, and organisational culture. Second, as a pragmatic quality improvement evaluation using an uncontrolled before-and-after design, assessment focused on process measures (processing time, occurrence of transcription-related errors in billing transfer, guidance duration, and completion rates) rather than direct clinical outcomes; the extent to which workflow improvement translates to longer-term health benefits was not assessed. In addition, variation over time was not formally analysed, and unmeasured contextual or secular changes may have contributed to some of the observed differences between fiscal years. A further limitation is that the completion-rate and guidance-duration measures were based on differently defined operational subgroups; the two denominators are therefore not directly comparable, and each measure should be interpreted as a separate process indicator rather than as part of a single nested cohort.

### Sustainability

Sustainability was supported by design choices that prioritised operational practicality over technical complexity. The workflow was built on widely available tools (a local relational database, a commercially available RPA platform, and a standard web-based form service), avoiding dependency on bespoke infrastructure or AI systems that may require specialist maintenance. By retaining paper letters as the primary communication method and offering the web form as an optional additional channel, the system preserved accessibility for diverse recipient groups.

Critically, the workflow was embedded in routine daily operations rather than implemented as a time-limited trial. Frontline ownership, in-house implementation capability, and an organisational culture receptive to incremental digitalisation supported continued use beyond the initial implementation period.

### Implications and transferability

The underlying principles described here may be transferable to other settings using different database or RPA tools, provided that workflow requirements are clearly defined and verification steps are preserved. These principles include centralising information management, standardising routine processes, separating human oversight from automated execution, and providing multiple response channels.

Transferability nonetheless depends on enabling conditions: the presence of a bridge role capable of translating frontline requirements into workable digital solutions, and an organisational culture that supports iterative, frontline-led improvement. Settings lacking these conditions may benefit from closer collaboration with information systems departments, external technical support, or staged capacity-building before pursuing comparable implementations.

## Conclusions

A non-AI-dependent integrated workflow using Microsoft Access, rule-based RPA, and an optional web form was associated with improvements in administrative efficiency and operational reliability in letter-based health guidance at a preventive medicine centre in Japan. Manual administrative handling time per case was substantially reduced, the completion rate improved, and the proportion of prolonged cases decreased. No transcription-related errors were detected during the post-implementation observation period. This pragmatic approach, which preserved human oversight and used widely available tools, may offer a useful model for process-level improvement in preventive health services. Multi-site evaluation and longer-term outcome assessment remain priorities for future work.

## Data Availability

Data are derived from routine operational records and are not publicly available due to institutional policies. Data may be available from the corresponding author on reasonable request and with appropriate approvals.

## Other Information

### Reporting guideline

This manuscript was prepared in accordance with the SQUIRE 2.0 (Standards for Quality Improvement Reporting Excellence) guidelines. A completed SQUIRE 2.0 checklist is provided as a supplementary file.^5^

### Funding

No specific funding was received for this work.

### Competing interests

The authors declare no competing interests.

## Acknowledgements

The authors thank Riho Sasaki, Satomi Kawakami, Ayaka Hagiwara, and Miku Yoshioka for their contributions to implementation support, data collection and operational work related to this quality improvement initiative.

## Author contributions

IK contributed substantially to implementation refinement, abstract development, and visualisation for academic presentation. EH conceived the original idea for the implementation and contributed substantially to its practical development in routine operations. MA contributed to implementation, data collection, and operational evaluation. RY contributed to critical revision of the manuscript, including language and clarity edits. YA supervised the project, drafted and revised the manuscript in English, and is the corresponding author. All authors critically reviewed the manuscript, approved the final version, and agree to be accountable for the work.

## Notes

### Competing Interest Statement

The authors have declared no competing interest.

### Author Declarations

The Yoshida Hospital Ethics Committee, Keiyukai Medical Corporation, gave ethical approval for this work on 21 April 2026. Approval number: 20260421001. Individual informed consent was waived by the ethics committee because this quality improvement study used aggregated, anonymised operational data and posed minimal risk to participants.

