## Supplementary Table S1 for "From Paper Letters to an Integrated Digital Workflow: Improving Efficiency, Reliability, and Engagement in Health Guidance"

**Supplementary Table S1. Guidance duration distribution**

This table provides the corrected distribution of guidance duration among cases where the programme was completed and the final evaluation was conducted by letter.

| Duration category | R4 / FY2022 (n=59) | R5 / FY2023 (n=158) |
|---|---|---|
| ≤160 days | 28 (47.5%) | 82 (51.9%) |
| 160–200 days | 13 (22.0%) | 43 (27.2%) |
| 200–240 days | 10 (17.0%) | 19 (12.0%) |
| ≥240 days | 8 (13.6%) | 14 (8.9%) |
| ≥200 days (200–240 + ≥240) | 18 (30.5%) | 33 (20.9%) |
| Mean duration (days) | 173 | 165 |

Denominator hierarchy (context):

FY2022: Eligible for intensive support n=756 to programme completed n=396 to final evaluation by letter n=59.

FY2023: Eligible for intensive support n=1,305 to programme completed n=674 to final evaluation by letter n=158.

Note: Duration was recalculated using the corrected method; prior values computed using an incorrect calculation period were excluded.
